# Vaccination reduces need for emergency care in breakthrough COVID-19 infections: A multicenter cohort study

**DOI:** 10.1101/2021.06.09.21258617

**Authors:** Amit Bahl, Steven Johnson, Gabriel Maine, Martha Hernandez Garcia, Srinivasa Nimmagadda, Lihua Qu, Nai-Wei Chen

## Abstract

**Importance:** While recent literature has shown the efficacy of the COVID-19 vaccine in preventing infection, its impact on need for emergency care/hospitalization in breakthrough infections remain unclear, particularly in regions with a high rate of variant viral strains.

**Objective:** We aimed to determine if vaccination reduces hospital visits and severe disease in breakthrough COVID-19 infections.

**Design:** Multicenter observational cohort analysis

**Setting:** Eight-hospital acute care regional health system in Michigan, USA

**Participants:** Consecutive adult patients with COVID-19 requiring emergency care (EC)/hospitalization were eligible participants. Between December 15, 2020 and April 30, 2021, 11,834 EC encounters with COVID-19 infection were included.

**Exposures:** COVID-19 vaccination

**Main Outcomes and Measures:** Primary endpoint was rate of COVID-19 emergency care/hospitalization encounters comparing unvaccinated (UV), partially vaccinated (PV), and fully vaccinated (FV) cases. Secondary outcome was severe disease represented as a composite outcome (ICU admission, mechanical ventilation, or in-hospital death).

Demographic and clinical variables were obtained from the electronic record. Vaccination data was obtained from the Michigan Care Improvement Registry and the Centers for Disease Control vaccine tracker.

**Results:** 10,880 (91.9%) UV, 825 (7%) PV, and 129 (1.1%) FV were included. Average age was 53.0 ± 18.2 and 52.8% were female. Accounting for the COVID-19 vaccination population groups in Michigan, the ED encounters/hospitalizations rate relevant to COVID-19 infection was 96% lower in FV versus UV (e^β^:0.04,95% CI 0.03 to 0.06, p <0.001) in negative binomial regression. COVID-19 EC visits rate peaked at 22.61, 12.88, and 1.29 visits per 100000 for the UV, PV, and FV groups, respectively. In the propensity-score matching weights analysis, FV had a lower risk of composite disease compared to UV but statistically insignificant (HR 0.84 95% CI 0.52 to 1.38).

**Conclusions:** The need for emergency care and/or hospitalization due to breakthrough COVID-19 is an exceedingly rare event in fully vaccinated patients. As vaccination has increased within our region, emergency visits amongst fully vaccinated individuals have remained low and occur much less frequently when compared to unvaccinated individuals. In cases of breakthrough COVID-19, if hospital-based treatment is required, elderly patients with significant comorbidities remain at high risk for severe outcomes regardless of vaccination status.

## Introduction

The COVID-19 pandemic has continued to cause significant morbidity and mortality around the globe with over 146 million cases and 3 million deaths as of April 25, 2021.^1^ In December of 2020, the FDA authorized emergency use of the Pfizer-BioNtech vaccine. It became the first of several vaccines to kick off the mass vaccination efforts across the United States.^2^ Subsequently, Moderna as well as Johnson and Johnson received emergency use authorization for their vaccines.^3^ While preliminary data from safety and efficacy trials have shown positive results, real-world data on its effectiveness is still lacking.^4^ Several small cohort studies and one large trial from Israel are currently our only insights into the actual rates of infection, hospitalization, and severe illness among vaccinated individuals.^5–7^Additionally, as COVID-19 variants emerge, we are in dire need of more data regarding the effectiveness of our current mass vaccination efforts.^8^

In-vitro studies have shown several variants and mutations to be more transmissible and less sensitive to natural or vaccine-induced antibodies compared to the wild-SARS CoV-2 virus.^8–10^ The Centers for Disease Control (CDC) has published a list of variants of concern and several of them include mutations in the spike protein incorporating the E484K and the L452R substitution.^9,11,12^ This is highly concerning, particularly in some regions in which new variant cases now outnumber the original wild-SARS CoV-2 strain.

In the latest surge of COVID-19, the state of Michigan has been more severely impacted than the rest of the United States.^1^ In Michigan the volume of peaked to over 7,000 new daily cases between April 5^th^ and April 12^th^ 2021.^13^ According to the CDC, over a 2 week period ending April 24, 2021, 10 COVID-19 variants were detected within the region.^14^ The most common, B.1.1.7 variant, has been identified as the cause of over 50% of new COVID-19 diagnosis in the State of Michigan.^14^ While the B.1.1.7 variant has shown to be associated with increased transmissibility, to date there has been no evidence to suggest or negate the impact on vaccine efficacy. However, in-vitro studies have noted a loss in neutralizing activity by vaccine-induced antibodies when the E484K mutation was introduced to the B.1.1.7 variant.^15^

Vaccination efforts in the State of Michigan have been ongoing since December.^13,16^ Given that approximately 42.72% of the state’s population was either partially or fully vaccinated as of April 30, 2021, it is unclear if immunization efforts have helped the situation in this recent COVID-19 surge in a population with a high incidence of variant strain disease.^13,14,16^ Therefore, we aim to evaluate if COVID-19 vaccination reduces rates of emergency care encounters and hospitalizations. Further we aim to understand vaccination impact on severe illness when breakthrough SARS-CoV-2 infection occurs.

## Methods

### Study Design, Setting and Participants

This was a multicenter observational cohort study through electronic health record (EHR; Epic Systems, Verona, WI) analysis to assess vaccine efficacy on need for emergency care/hospitalizations and severe outcomes in patients with breakthrough COVID-19 infection comparing fully vaccinated (FV), partially vaccinated (PV), and unvaccinated (UV) patients. The study was conducted at Beaumont Health, an eight-hospital acute care regional health system caring for 2.2 million people across the communities within the Metro Detroit area. The hospitals range from a large tertiary care academic center to intermediate-sized and smaller community hospitals.

Consecutive adult patients greater than 18 years of age presenting to the emergency department with confirmed COVID-19 as a primary diagnosis were eligible for inclusion. Patients with prior laboratory confirmed COVID-19 infection, pediatric patients, or those still hospitalized after the designated follow-up date of May 15, 2021 were excluded. The study was approved by the Institutional Review Board at Beaumont Health and registered on clinicaltrials.gov (Identifier: NCT04912700). Written informed consent requirement was waived due to the retrospective nature of this study. Data were analyzed and interpreted by the authors.

### Study Definitions

Patients were categorized as either UV, PV, or FV. UV individuals were defined as having positive laboratory COVID-19 testing with no record of immunization against COVID-19 or first-dose vaccination after symptom onset. PV individuals were defined as having positive laboratory COVID-19 testing and symptom onset after a single dose of either mRNA (Pfizer, Moderna) vaccine, or < 14 days after the second dose of either mRNA vaccine (Pfizer, Moderna) or < 14 days after the administration of the single dose of viral vector vaccine (Johnson & Johnson). FV individuals were defined as having positive laboratory testing for COVID-19 and symptom onset >14 days since administered of second dose of either mRNA vaccine, or >14 days since administration of viral vector vaccine (Johnson & Johnson).

COVID-19 infection was defined as primary diagnosis of COVID-19 by ICD-10-CM codes in the EHR and either laboratory confirmed positive result (rapid antigen testing or reverse-transcriptase-polymerase-chain-reaction (RT-PCR) by nasopharyngeal swab) or reference to confirmed laboratory diagnosis in the emergency encounter provider note.

### Data sources/measurement

EHR data was used to confirm COVID-19 infection and categorize vaccinated patients. For patients with primary diagnosis of COVID-19 without laboratory confirmed COVID diagnosis within the institutional EHR, emergency care provider records were manually reviewed by two independent physicians to confirm diagnosis of infection. Physicians also reviewed provider notes for all PV and FV patients to determine onset of symptoms. The date of symptom onset was used to accurately categorize UV, PV, and FV groups.

Demographic, clinical, and outcomes data were obtained from the EHR. Demographics included age, race, and gender. Clinical data included comorbidities, body mass index (BMI), and number of previous ED visits within the past 6 months. ICD-10-CM codes for comorbidities were used to calculate the Elixhauser comorbidity weighted scores as described by the Agency of Healthcare Research and Quality (AHRQ).^17^ Hospital clinical data included level of care required, extracorporeal membrane oxygenation (ECMO), renal replacement therapy, type of oxygen or ventilation therapy, and need for vasopressors.

Hospital admission was based on the clinical judgment of the treating emergency medicine provider and length of stay was calculated for all admitted patients. Discharge disposition post-hospitalization was based on patients’ clinical condition. Patients were either discharged to home, skilled nursing home, rehabilitation facility, hospice, or expired in the hospital.

Vaccination data was made available by the state of Michigan via the Michigan Care Improvement Registry (MCIR) and therefore captured patients who had been vaccinated outside of the Beaumont Health system.^13^ This data included vaccine type as well as date of administration. Vaccination prevalence across the population of Michigan was captured weekly via the Centers of Disease Control state-specific vaccine tracker.^14^

### Outcome Measures

The primary outcome was rate of COVID-19 emergency care/hospitalization encounters among UV, PV, and FV groups.

Secondary outcomes included severe disease represented as a composite outcome (ICU admission, mechanical ventilation, or in-hospital death), hospital length of stay, renal replacement therapy, ECMO supplemental oxygen (none, low flow therapy, and high flow therapy), and noninvasive ventilation.

### Statistical Analysis

Bivariate analyses were stratified by vaccinated status (UV, PV, FV) using means ± standard deviations and medians with interquartile ranges (IQRs) for continuous variables and frequencies with percentages for categorical variables. Kruskal-Wallis (exact) test (continuous variables) and Chi-squared or Fisher’s exact test (categorical variables) were used to compare differences among three categories of vaccinated status.

To investigate the effect of vaccination status on the occurrence of COVID-19 emergency care/hospitalization encounters, Negative Binomial regression analysis with the log-link, accounting for any potential overdispersion, was used based on weekly rates per 100000 to the State COVID-19 vaccination population groups. To characterize variation in weekly rates across the study period, joinpoint regression analyses (ie, segmented trend analysis with continuity constraint) by vaccination status on the log-scale with up to 2 joinpoints were used through grid search of joinpoints by Monte Carlo permutation tests.^18^

Cox proportional hazards regression models were used to examine the association between vaccination status and severity of illness, a composite outcome of ICU admission, mechanical ventilation, or in-hospital death. An initial multivariable Cox regression model was built, controlling for demographic characteristics and clinical variables including the Elixhauser weighted score and occurrence of ED visits prior to 6 months. We applied the test for proportionality assumption based on the Schoenfeld residuals. Stratified Cox regression was applied to adjust the potential nonproportional hazards. In addition, to eliminate the bias of patient characteristics on exposure of vaccination status, we used propensity-score methods to reduce the effect of confounding. The individual propensities for exposure of vaccination status were estimated from a multivariable multinomial logistic regression model that included the same covariates as the multivariable Cox regression. A three-way 1:1:1 nearest neighbor matching was conducted for evaluating the effect of vaccination on severity of illness.

Furthermore, in settings with rare outcomes and unequal exposure distributions of vaccination status, we also applied matching weights for assessing the association.^19^ The covariate balance was achieved after matching or weighting (results not shown). All tests of statistical significance were indicated with two-sided 95% confidence intervals (CIs) or p < 0.05. Analyses were performed using Joinpoint Regression Program v4.7.0.0, R-4.0.2 (R Foundation for Statistical Computing), and SAS v9.4 (SAS Institute, Inc., Cary, NC).

## Results

Between December 15, 2020 and April 30, 2021 there were a total of 169000 ED encounters within our hospital system. We identified 11895 of these encounters that met our inclusion criteria. After further exclusion of 61 encounters, that remained admitted after our designated follow up date, we were able to analyze 10880 unvaccinated, 825 partially vaccinated, and 129 fully vaccinated ED encounters. (Figure 1).

**Figure 1.**
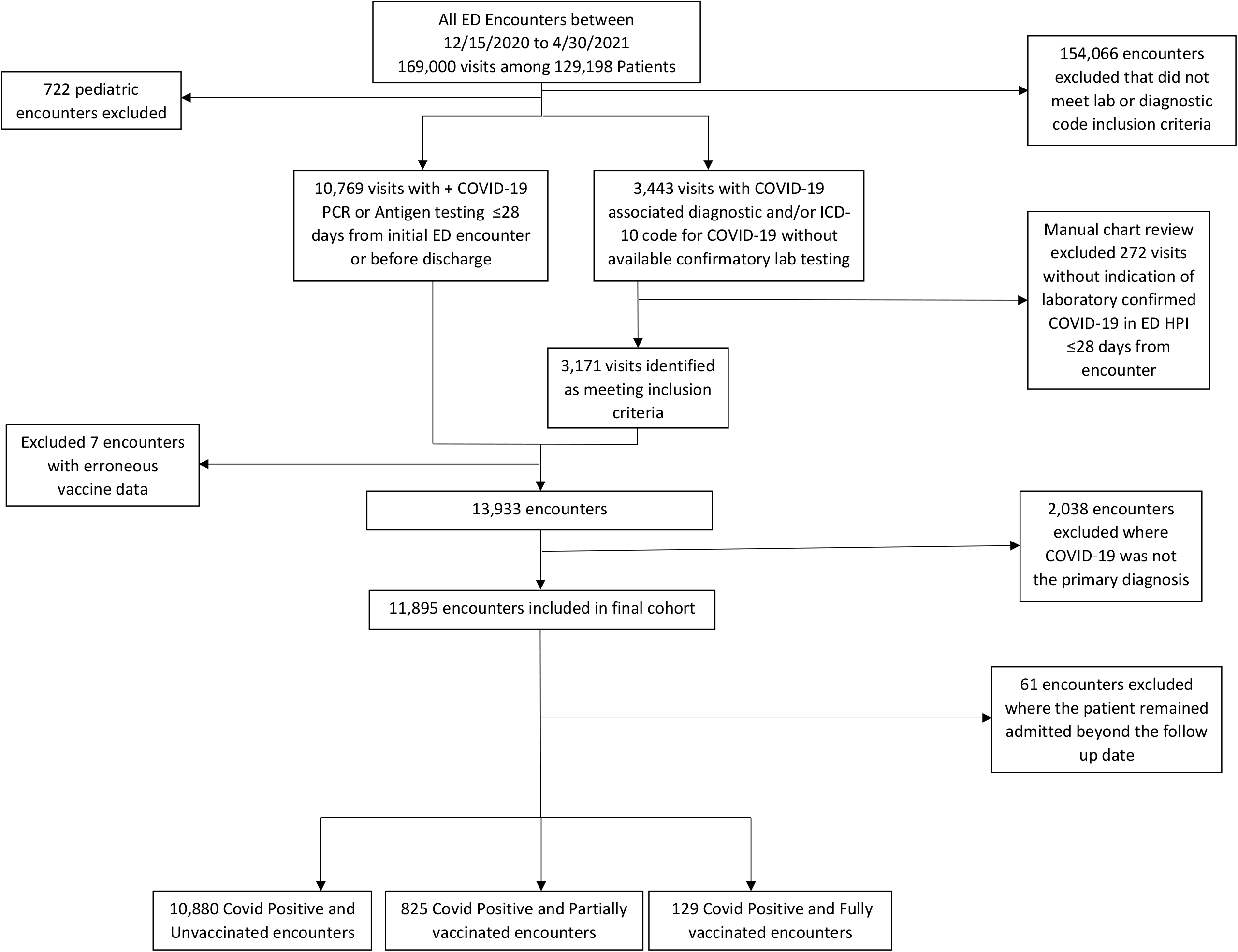
Screening and Categorization of all hospital-based COVID-19 cases into UV, PV, and FV groups Patients with COVID-19 infection with an ED encounter were eligible participants. Patients with secondary diagnosis of COVID-19 and age less than 18 years were excluded. Patients with primary COVID-19 diagnosis without reference to confirmed testing in the emergency provider note were also excluded. Included patients were then categorized into UV, PV, and FV cohorts. UV individuals had no record of immunization against COVID-19 or first-dose vaccination after symptom onset. PV individuals had symptom onset after a single dose of either mRNA (Pfizer, Moderna) vaccine, or < 14 days after the second dose of either mRNA vaccine (Pfizer, Moderna) or < 14 days after the administration of the single dose of viral vector vaccine (Johnson & Johnson). FV individuals had symptom onset >14 days since administered of second dose of either mRNA vaccine, or >14 days since administration of viral vector vaccine (Johnson & Johnson).

Complete demographic and comorbidity data for the cohort is displayed in Table 1. There were differences among groups in age, race, ED visits in the prior 6 months, and the Elixhauser comorbidity weighted score. The average ages were 52.1 ± 18.2, 62.5 ± 15.3, and 70.3 ± 16.4 (p< 0.001) for the UV, PV, and FV groups, respectively. There was a larger proportion of African American patients in the UV group at 3452 (31.7%) vs 198 (24%) and 13 (10%) in the PV and FV groups, respectively. The FV group had a statistically higher number of encounters with repeat ED visits within the prior 6 months at 48 (37.2%), vs 196 (23.8%) and 2292 (21.1%) (p< 0.001) in the PV and UV groups, respectively. The average Elixhauser comorbidity weighted scores were 4.3 ± 8.8, 6.7 ± 9.6, and 10.3 ± 11.1 (p< 0.001) in the UV, PV, and FV groups, respectively.

**Table 1.** Patient characteristics by vaccination status for all ED visits

| Variables<sup>‡</sup> | All | Vaccination Status |  |  | *p* value |
| --- | --- | --- | --- | --- | --- |
|  |  | Unvaccinated | Partially Vaccinated | Fully Vaccinated |  |
| n | 11834 | 10880(91.9) | 825 (7.0) | 129(1.1) |  |
| Age, years | 53.0 ± 18.2 | 52.1 ± 18.2 | 62.5 ± 15.3 | 70.3 ± 16.4 | < 0.001 |
|  | 53.0 (39.0, 66.0) | 52.0 (38.0, 65.0) | 63.0 (52.0, 74.0) | 72.0 (62.0, 82.0) |  |
| 18 to 40- | 3053 (25.8) | 2983 (27.4) | 63 (7.6) | 7 (5.4) | < 0.001 |
| 40 to 65- | 5542 (46.8) | 5126 (47.1) | 386 (46.8) | 30(23.3) |  |
| ≥ 65 | 3239 (27.4) | 2771 (25.5) | 376 (45.6) | 92(71.3) |  |
| Sex |  |  |  |  |  |
| Male | 5590 (47.2) | 5130 (47.2) | 400 (48.5) | 60(46.5) | 0.75 |
| Female | 6244 (52.8) | 5750 (52.8) | 425 (51.5) | 69(53.5) |  |
| Race |  |  |  |  |  |
| White/Caucasian | 7134 (60.3) | 6467 (59.4) | 559 (67.8) | 108(83.7) | < 0.001 |
| Black/African American | 3663 (30.9) | 3452 (31.7) | 198 (24.0) | 13(10.1) |  |
| Other | 1037 (8.8) | 961 (8.8) | 68 (8.2) | 8(6.2) |  |
| BMI, kg/m<sup>2</sup> | 32.0 ± 8.6 | 32.1 ± 8.7 | 32.1 ± 7.9 | 30.1 ± 8.4 | 0.01 |
|  | 31.0 (26.3, 36.2) | 31.0 (26.3, 36.2) | 31.1 (26.5, 36.8) | 28.1 (23.7, 34.1) |  |
| < 30 | 5340 (45.1) | 4898 (45.0) | 369 (44.7) | 73(56.6) | 0.03 |
| ≥ 30 | 6494 (54.9) | 5982 (55.0) | 456 (55.3) | 56(43.4) |  |
| Elixhauser weighted score | 4.5 ± 8.9 | 4.3 ± 8.8 | 6.7 ± 9.6 | 10.3 ± 11.1 | < 0.001 |
|  | 0.0 (0.0, 10.0) | 0.0 (0.0, 9.0) | 5.0 (0.0, 13.0) | 8.0 (2.0, 16.0) |  |
| < 0 | 2687 (22.7) | 2492 (22.9) | 178 (21.6) | 17(13.2) | < 0.001 |
| 0 to 10 | 6538 (55.3) | 6099 (56.1) | 384 (46.5) | 55(42.6) |  |
| > 10 | 2609 (22.0) | 2289 (21.0) | 263 (31.9) | 57(44.2) |  |
| ED visits prior to 6 months |  |  |  |  |  |
| No | 9298 (78.6) | 8588 (78.9) | 629 (76.2) | 81(62.8) | < 0.001 |
| Yes | 2536 (21.4) | 2292 (21.1) | 196 (23.8) | 48(37.2) |  |

Abbreviations: ED=emergency department; BMI=body mass index.
<sup>‡</sup> For continuous variables, means ± standard deviations and medians (interquartile ranges, IQRs) were presented. For categorical variables, frequencies and percentages within parentheses were presented. Missing BMI were less than 5% of observations and imputed by mean.

Patients that experienced severe disease inclusive of composite ICU admission, mechanical ventilation, or in-hospital mortality, had similar baseline characteristics displayed in Table 2. Our composite outcome occurred in 733 (6.8% of 10880) encounters in the UV group, 85 (10.3% of 825) encounters in the PV group, and 16 (12.4% of 129) in the FV group. Among all groups there were 442 (3.7% of 11834) deaths. For each group, death occurred in 384 (3.5% of 10880) UV patients, 50 (6.1% of 825) PV patients, and 8 (6.2% of 129) FV patients.

**Table 2.**
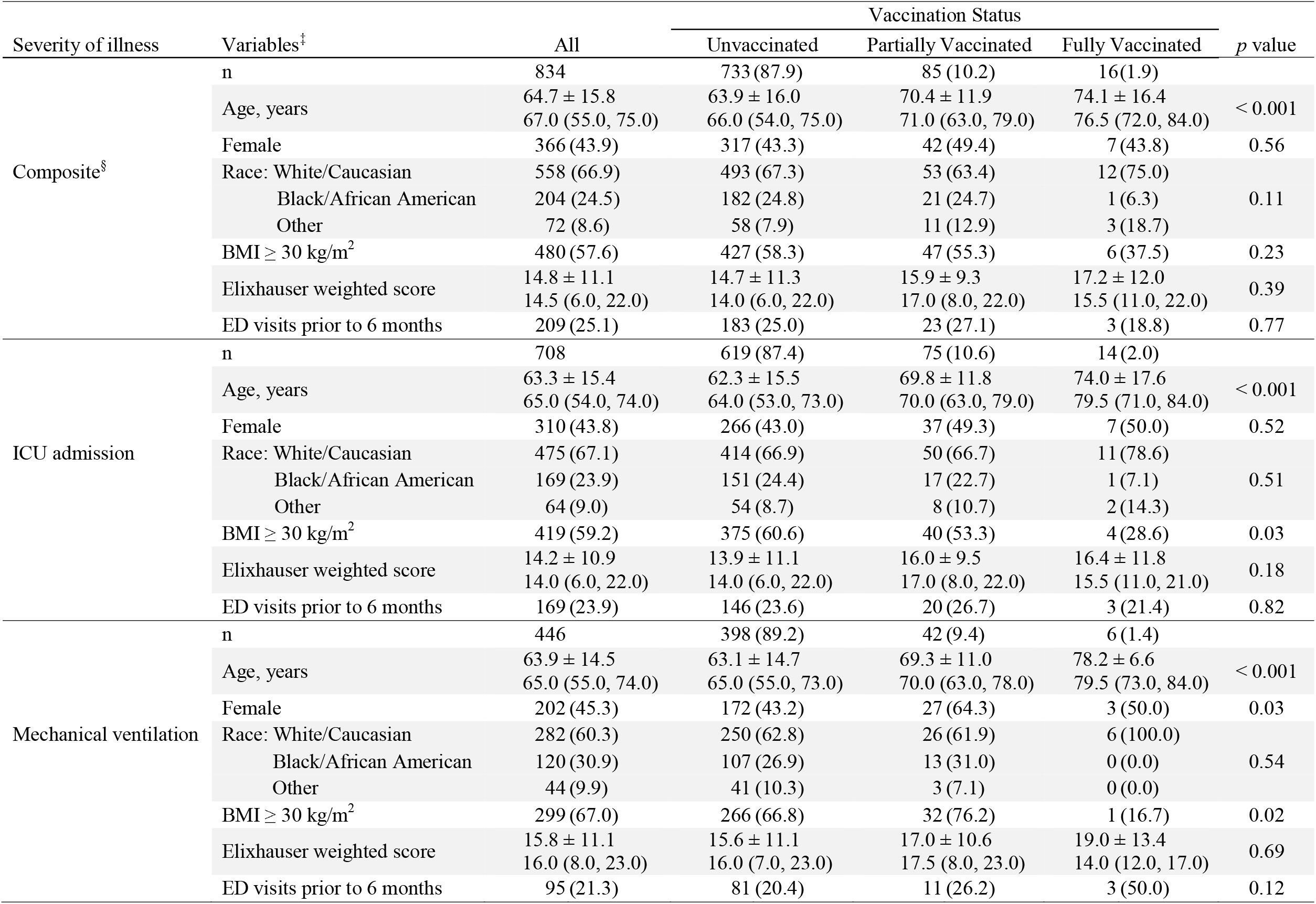
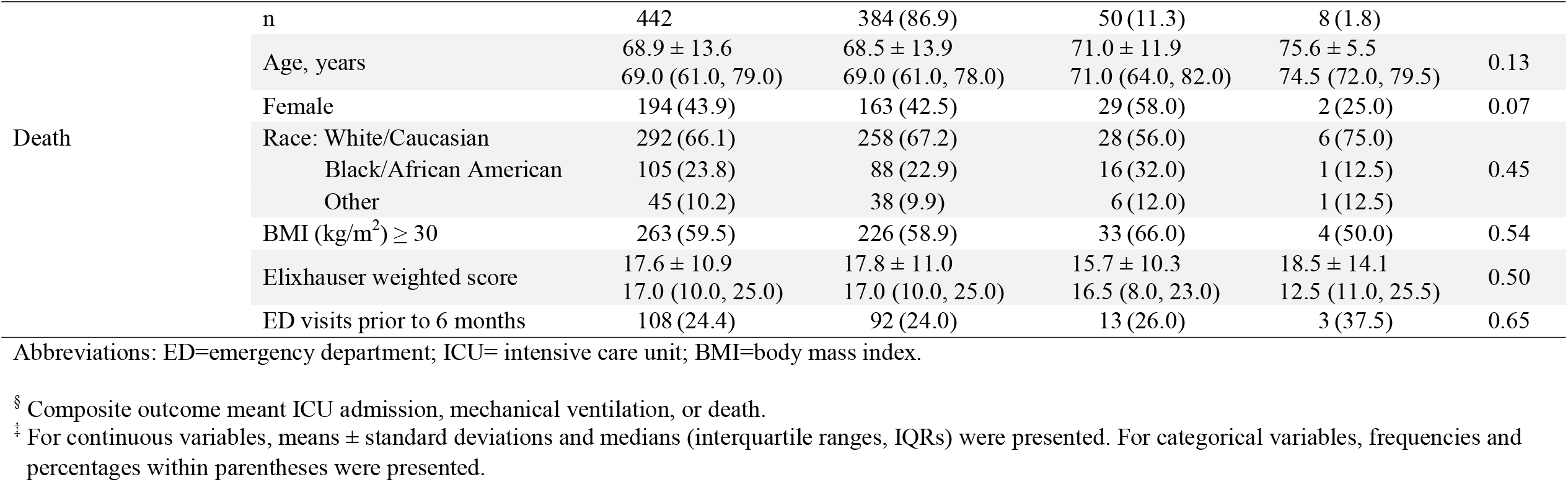
Patient characteristics by vaccination status on outcomes of severity of illness for all ED visits

We evaluated the effect of vaccination status on the weekly rate of COVID-19 emergency care/hospitalization encounters to the State COVID-19 vaccination population groups in negative binomial model. Table 3 demonstrates on average, a significant 96% lower rate of COVID-19 ED visits in the FV group compared to UV group (e^β^: 0.04, 95% CI 0.03 to 0.06, p<0.001). Figure 2 plots the rate of COVID-19 ED visits to the State COVID-19 vaccination population groups across study period for each category of vaccinated status. The peak rate of COVID-19 ED visits per 100000 occurred between 4/4/21 and 4/17/21 for all three groups. The crude rate peaked at 22.61, 12.88, and 1.29 visits per 100000 for the UV, PV, and FV groups, respectively. During the increase in COVID-19 presenting to the ED between 2/21 and 4/21, the rate of visits for the fully vaccinated group oscillated between 0.00 to 1.29 per 100000. During this same spike, the rate of visits for unvaccinated individuals went from 1.97 up to 22.61 per 100000.

**Table 3.** Results of effect of vaccination status on rate of COVID-19 ED visits

| Effects<sup>§</sup> | | | Estimate, $\beta$, (SE) | $e^{\beta}$ (95% CI) | *p* value |
| --- | --- | --- | --- | --- | --- |
| Fully vaccinated | vs. | Unvaccinated | −3.20 (0.23) | 0.04 (0.03, 0.06) | < 0.001 |
| Fully vaccinated | vs. | Partially vaccinated | −2.55 (0.22) | 0.08 (0.05, 0.12) | < 0.001 |
| Partially vaccinated | vs. | Unvaccinated | −0.65 (0.18) | 0.52 (0.37, 0.74) | 0.001 |

Abbreviations: SE=standard error; CI=confidence interval.
<sup>§</sup> Negative Binomial regression analyses with the log-link were based on weekly rates per 100000 to the State COVID-19 vaccination population groups.

**Figure 2.**
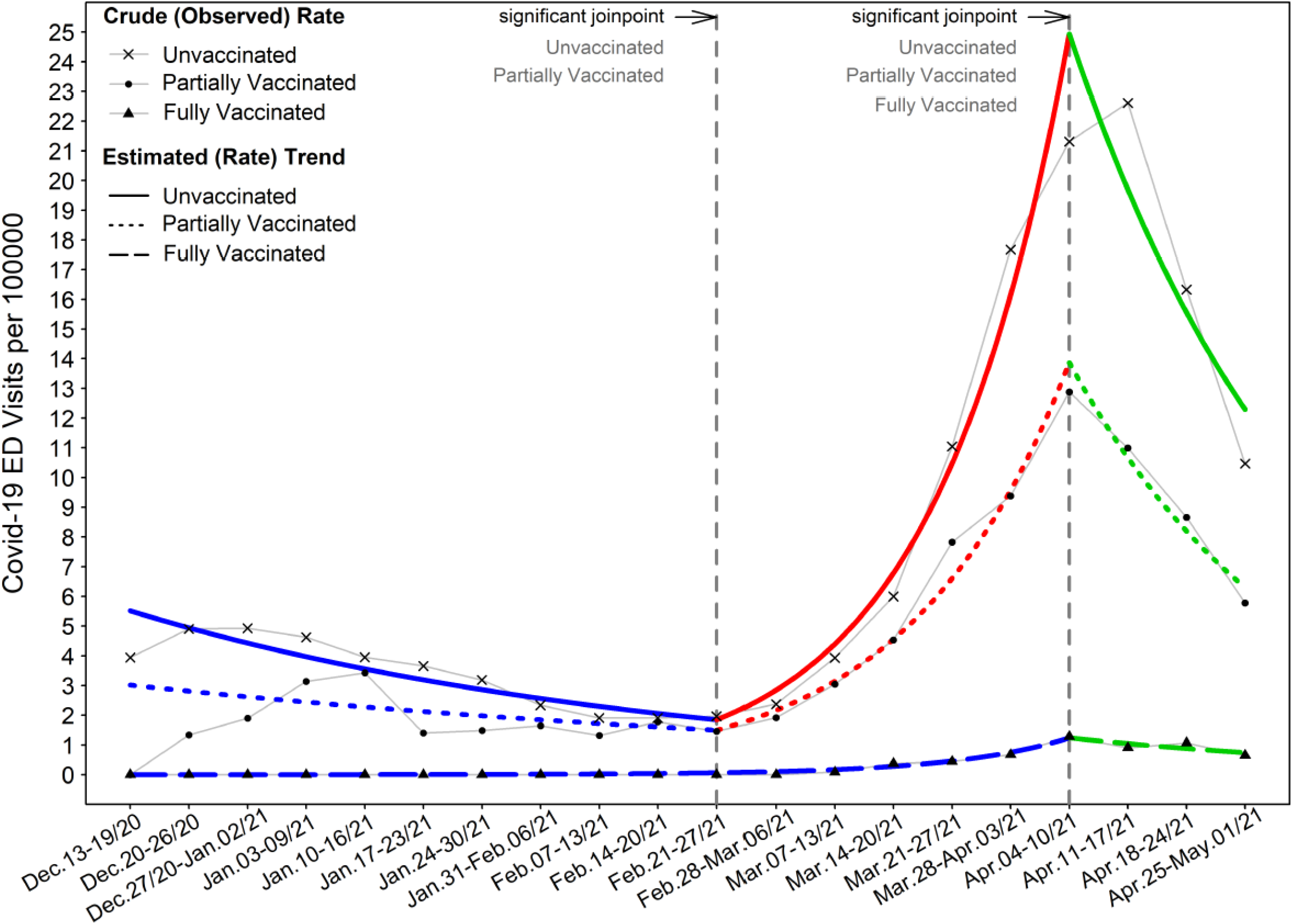
ED encounters of COVID-19 patients among UV, PV, and FV groups Results shown are for the entire study cohort of adult COVID-19 patients presenting from December 15, 2020 thru April 30, 2021. Case rate of emergency encounters proportionated to the State COVID-19 vaccination population groups. Weekly crude and estimated trend of COVID-19 infection ED visits for each vaccinated group are depicted as number of cases per 100000 over study period. The line graph illustrates the estimated trend of infection ED encounters (visits) for each vaccinated group. When the state FV population size was only 19 individuals between 12/27/2020 and 1/2/2021, one ED visit occurred in FV group which was not included in analysis due to the bias of an extreme outlier in trend analysis.

We examined the association of vaccination status on severe composite disease. In the propensity-score matching weights analysis, results indicate that compared to UV group, FV group had a lower risk of severe composite disease but statistically not significant (hazard ratio HR 0.84, 95% CI 0.52 to1.38); partially vaccinated group was not associated with a significantly higher or lower risk of severe composite disease (HR 1.03, 95% CI 0.78 to 1.35). Propensity-score three-way matching analysis yielded similar results. (Table 4)

**Table 4.**
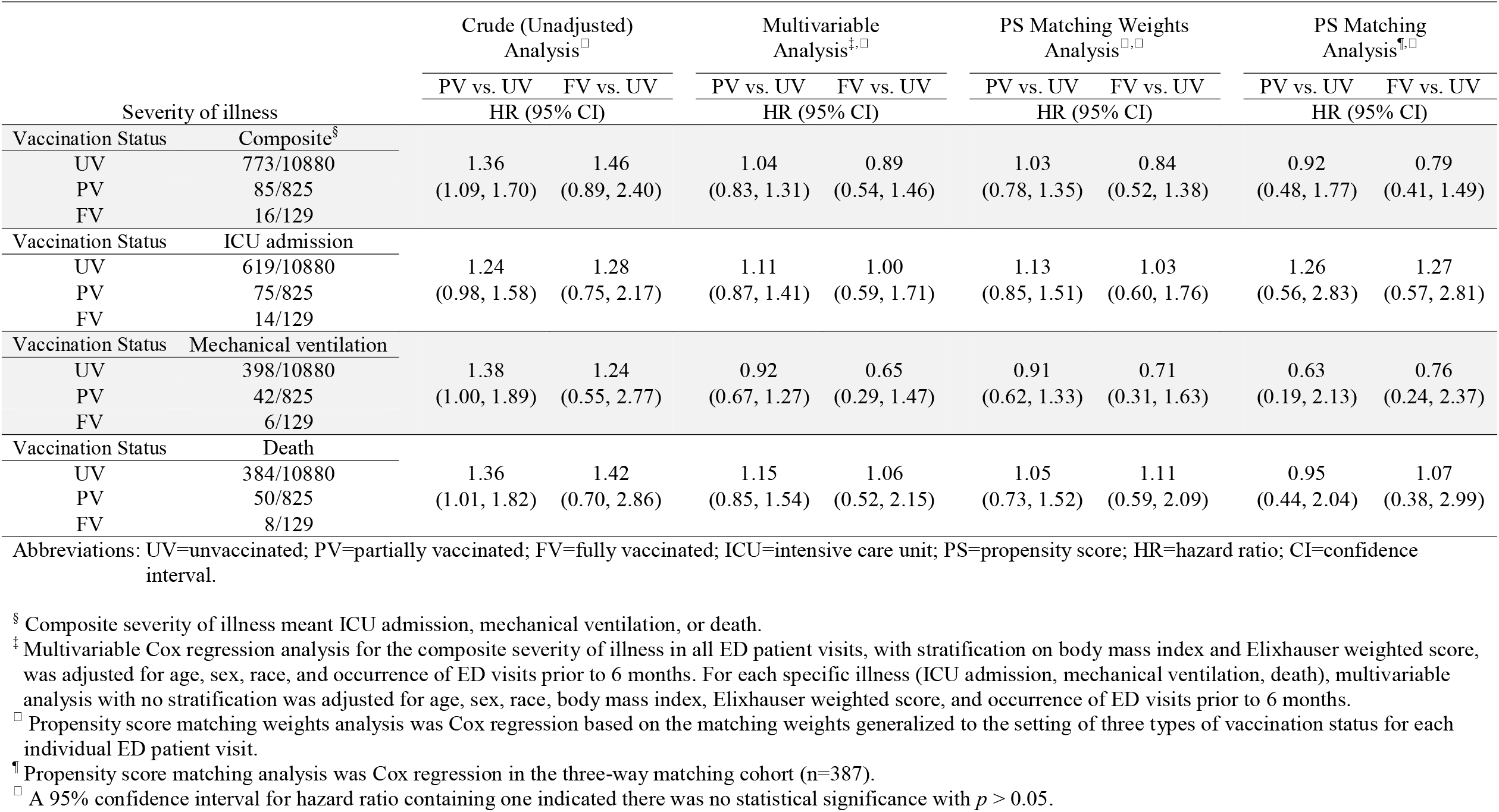
Association between vaccination status and severity of illness

Use of ECMO was only seen amongst 4 patients in the UV group with zero instances in the PV and FV groups. The remainder of secondary outcomes by group type are displayed in supplemental Table 1.

## Discussion

Despite aggressive vaccination efforts in Michigan, the rapid increase in new cases during our study period highlights the need to quantify the benefit of these efforts. This study demonstrated that regardless of the high incidence of daily COVID-19 infections, with a majority due to variant strains, fully vaccinated individuals remained substantially less likely to seek emergency care or become hospitalized. ^13,14^ Compared to unvaccinated cases, significantly fewer fully vaccinated patients with breakthrough COVID-19 infection required emergency care and/or hospitalization. Notably, despite the surge of COVID-19 cases during the week of April 4^th^ 2021 the rate of COVID-19 related emergency care for fully vaccinated patients remained low. While this study did not specifically assess for efficacy of the vaccination in preventing disease in the community, this study addressed a possibly more relevant clinical question of likelihood for breakthrough COVID-19 infection to require hospital-based treatment.

Our cohort of fully vaccinated patients with breakthrough COVID-19 infections comprised only 1% of COVID-19 emergency care visits during the study period. Within this group, we found that those who required hospitalization and developed severe illness were geriatric patients. Not surprisingly, similar to other vaccinations with reduced effectiveness in the elderly population, this geriatric group represented the population most at risk for serious adverse outcomes.^20,21^ Each of the three study groups included patients as young as 19 years of age. In the fully vaccinated group, all 8 deaths and 6 intubations occurred in patients over the age of 65. While in the unvaccinated group, patients as young as 21 died while hospitalized and patients as young as 19 required mechanical ventilation.

Accompanied with advancing age, chronic disease burden was an important contributing factor to the adverse outcomes in fully vaccinated patients. With an average baseline elixhauser score >10, this group was at high-risk for near term death. Existing literature suggests that a weighted score of 10 predicts a slightly less than 10% risk of in hospital death, while a score of approximately 37 predicts a 50% chance of death while admitted to the hospital. ^22^ In this fragile group, risk of in-hospital death was similar to the matched unvaccinated group. While the mortality rate from COVID-19 infection has declined from the beginning of the pandemic in which nearly 30% of hospitalized patients died, the death rate was still 6.2% in the fully vaccinated group in our cohort. ^23–25^ While this mortality rate is concerning, it is important to understand this outcome in the context of other similar disease processes. For example, comparatively, other endemic respiratory viral illnesses such as influenza which can cause severe disease requiring ICU admission in up to 10% of cases that require hospitalization and mortality rates up to 8.3%. ^26,27^

It is unclear if the vaccination results will hold steady with ongoing viral mutations and emergence of viral variants. Some data suggests that viral mutations may reduce the efficacy of vaccination. For instance, Collier et al. observed a loss in neutralizing activity by vaccine-induced antibodies when the E484K mutation was introduced to the B.1.1.7 variant. This may lead to the need of a substantially larger amount of antibodies to prevent infections.^15^ It is also unknown if protective effect of immunization regarding severe disease will wane and expose vulnerable groups to more severe disease. However, our study demonstrated that for now, with over 50% prevalence of variant disease in the region, vaccination is likely efficacious against existing variants as the rate of breakthrough infections requiring hospital treatment in fully vaccinated patients was low.

Our study had some limitations. The observational cohort study design was a limitation and it is possible that some patients with COVID-19 were not included despite our careful screening of all diagnostic test types. Further, patients with potential COVID-19 with negative laboratory testing were not included in this analysis. As high-risk patients often receive multiple tests to rule out infection, the miss rate was likely small. Another limitation was reliance of electronic health record data. The data is reliant on accurate documentation and it is likely that some input errors occurred. Further, some patients had incomplete data and this limited our analysis. Selection bias was another limitation as patients that were still hospitalized after the cutoff follow-up date (May 15) were excluded from the analysis. Fortunately, only 61 (0.5%) patients were still hospitalized. While some of the potentially more severe cases with longer hospital durations were excluded from the analysis, only three cases were excluded due to this reason in the fully vaccinated group, the main population of interest. As the information was time sensitive, we decided it was appropriate to move forward with analysis before waiting for all patient encounters to be complete. Additionally, we assumed vaccination rate of study patients was similar to the published data regarding vaccination status from the state population. It is possible that there were slight variations that were not captured with this methodology. We also assumed our study population had similar rates of variant disease as reported by the state. Test samples were periodically sent to and audited by the state laboratory and internal hospital quality data confirmed our assumptions on rates of variants. Finally, while vaccination data from the state registry was generally robust, in seven cases the data from the MCIR was insensible with some patients receiving a combination of vaccination types or more than two doses. These cases were excluded from the analysis. Additionally, we could not make any vaccination specific conclusions. As the number of fully vaccinated patients was small in our cohort, further sub-analysis by vaccination type was not possible.

In summary, emergency visits and hospitalization in fully vaccinated patients with breakthrough COVID-19 are extremely rare events even in a region with high incidence of variants. When hospitalization occurs, immunized patients are older with many comorbidities. In this high-risk population, risk for severe disease was similar in unvaccinated and vaccinated patients. Future studies are needed to reassess vaccination efficacy broadly and by type of vaccine as mutations and variants evolve.

## Data Availability

The datasets generated during and/or analyzed during this study are available from the corresponding author upon reasonable request.

---

**Supplemental Table 1.** Treatments and clinical outcomes by vaccination status for hospitalized patients

| Variables<sup>‡</sup> | All | Vaccination Status |  |  | *p* value |
| --- | --- | --- | --- | --- | --- |
|  |  | Unvaccinated | Partially Vaccinated | Fully Vaccinated |  |
| n | 5860 | 5250(89.6) | 515 (8.8) | 95(1.6) |  |
| **Treatments** |  |  |  |  |  |
| Any oxygen therapy | 4502 (76.8) | 4042 (77.0) | 396 (77.0) | 64(67.4) | 0.09 |
| Nasal cannula | 2832 (48.3) | 2563 (48.8) | 231 (44.8) | 38(40.0) | 0.06 |
| High flow oxygen | 733 (12.5) | 656 (12.5) | 67 (13.0) | 10(10.5) | 0.79 |
| Non-invasive ventilation | 494 (8.4) | 428 (8.2) | 56 (10.9) | 10(10.5) | 0.08 |
| Mechanical ventilation | 443 (7.6) | 395 (7.5) | 42 (8.2) | 6(6.3) | 0.79 |
| ECMO | 4 (0.1) | 4 (0.1) | 0 (0.0) | 0(0.0) | 1.00 |
| Renal replacement therapy | 144 (2.5) | 130 (2.5) | 14 (2.7) | 0(0.0) | 0.28 |
| Vasopressors | 399 (6.8) | 348 (6.6) | 45 (8.7) | 6(6.3) | 0.19 |
| **Outcomes** |  |  |  |  |  |
| Hospital length of stay, days | 7.2 ± 6.9<br>5.2 (3.1, 8.5) | 7.2 ± 7.0<br>5.2 (3.1, 8.4) | 7.3 ± 6.3<br>5.4 (3.4, 9.0) | 7.0 ± 5.2<br>6.1 (3.1, 9.4) | 0.27 |
| Hospital disposition |  |  |  |  |  |
| Home | 4539 (77.5) | 4110 (78.3) | 368 (71.5) | 61 (64.2) | < 0.001 |
| Rehabilitation facilities | 79 (1.4) | 72 (1.4) | 7 (1.4) | 0(0.0) |  |
| Skilled nursing home | 600 (10.2) | 514 (9.8) | 66 (12.8) | 20(21.1) |  |
| Hospice | 152 (2.6) | 125 (2.4) | 21 (4.1) | 6(6.3) |  |
| Transferred | 54 (0.9) | 50 (0.9) | 4 (0.8) | 0(0.0) |  |
| Death | 436 (7.4) | 379 (7.2) | 49 (9.5) | 8(8.4) |  |

Abbreviations: ECMO=extracorporeal membrane oxygenation.
<sup>‡</sup> For continuous variables, means ± standard deviations and medians (interquartile ranges, IQRs) were presented. For categorical variables, frequencies and percentages within parentheses were presented.

